# Robustness of Deep Learning Segmentation Across Heterogeneous Uterine MRI Datasets: A Multi-Task Comparison of U-Net, Swin-UNETR, MedNeXt, and nnU-Net

**DOI:** 10.64898/2026.07.22.26358583

**Authors:** Daniel A. Di Giovanni, Akiyo Takada, Evan McNabb, Jérémy Dana, Hajime Yokota, Takahiro Tsuboyama, Rita Zakarian, Martin Vallières, James Man Git Tsui, Caroline Reinhold

**Affiliations:** Department of Diagnostic Radiology, McGill University, Montreal, QC, Canada; Augmented Intelligence & Precision Health Laboratory (AIPHL), Research Institute of the McGill University Health Centre, Montreal, QC, Canada; The Research Institute of the McGill University Health Centre, Montreal, QC, Canada; Diagnostic Radiology and Radiation Oncology, Chiba University Graduate School of Medicine, Chiba, Japan; Medical Imaging, McGill University Health Centre, Montreal, QC, Canada; Medical Physics Unit, Department of Oncology, Faculty of Medicine and Health Sciences, McGill University, Montreal, QC, Canada; Department of Radiology, Centre hospitalier de l’Université de Montréal (CHUM), Montréal, QC, Canada; Centre de recherche du Centre hospitalier de l’Université de Montréal (CRCHUM), Montréal, QC, Canada; Department of Radiology, The University of Osaka Graduate School of Medicine, Osaka, Japan; Department of Radiation Oncology, McGill University, Montreal, QC, Canada

**Keywords:** Uterine MRI, Medical image segmentation, Domain shift, Robustness, nnU-Net, MedNeXt

## Abstract

Reliable medical image segmentation requires performance that is stable across tasks, acquisition settings, and evaluation domains. We evaluated U-Net, Swin-UNETR, and MedNeXt as manually configured architectures and nnU-Net as a self-configuring reference across three T2-weighted uterine MRI datasets: public multiclass uterine anatomy and fibroid segmentation (UMD, n=300), institutional endometrial cancer tumor segmentation (n=206), and institutional uterine mass lesion segmentation (n=234). A separate UMD-style cohort (n=12), relabeled to the same annotation ontology, was used to assess external domain shift. Fixed data partitions, fold ensembling, and matched evaluation were used across models. Performance was assessed with Dice, 95th-percentile Hausdorff distance, average symmetric surface distance, absolute volume difference, and paired bootstrap comparisons with within-dataset Holm correction. MedNeXt was the strongest manually configured architecture, whereas nnU-Net achieved the highest Dice on all internal datasets: 0.761 for UMD, 0.746 for endometrial cancer, and 0.814 for uterine mass segmentation, compared with 0.722, 0.726, and 0.789 for MedNeXt. The nnU-Net-MedNeXt separation was largest for multiclass UMD segmentation and smaller for the binary tasks. All models degraded substantially on external testing; Dice was 0.542 for nnU-Net, 0.490 for MedNeXt, 0.396 for U-Net, and 0.287 for Swin-UNETR. These findings show that architecture, task definition, and dataset-adaptive pipeline configuration jointly influence uterine MRI segmentation, while external domain shift remains a major source of failure.

## 1. Introduction

Accurate medical image segmentation underpins quantitative imaging workflows, including phenotyping, therapy planning, and imaging-derived biomarker development. While substantial progress has been achieved in segmentation performance on well-studied organs, these advances do not necessarily translate to anatomically and pathologically complex targets. The uterus represents a particularly challenging and clinically important case: its zonal anatomy on T2-weighted MRI, including the endometrium, junctional zone, and myometrium, varies with physiology, is dynamically mobile within the pelvis, and is frequently distorted by both benign and malignant disease [1–3]. These characteristics make reliable segmentation essential for consistent quantification, yet difficult to achieve across diverse clinical scenarios.

Uterine leiomyomas (LMs), or fibroids, benign smooth muscle tumors affecting up to 25-77% of women, are highly prevalent and represent a major source of morbidity, motivating imaging-based mapping of lesion burden and location for clinical reporting and procedural planning [4–6]. In contrast, uterine leiomyosarcoma (uLMS) is a rare but aggressive malignancy, accounting for approximately 1-2% of uterine cancers, yet posing substantial diagnostic and management challenges [7]. Critically, a subset of LMs demonstrate atypical imaging features that overlap with uLMS, resulting in persistent diagnostic uncertainty even on high-quality MRI. This overlap is clinically consequential, as uLMS often presents with non-specific features and is frequently diagnosed only after surgical intervention [8]. This distinction has direct management implications: presumed benign fibroids may be treated with minimally invasive approaches, whereas suspected uLMS generally requires *en bloc* resection to support margin assessment and reduce the risk of tumor dissemination. MRI, the modality of choice for uterine mass characterization due to its superior soft-tissue contrast and multiparametric capability, plays a central role in this setting [9]. However, the variability in appearance across benign and malignant lesions combined with overlapping imaging phenotypes creates a challenging environment for both human interpretation and automated analysis. Despite this clinical importance, segmentation of uterine anatomy and uterine lesions remains largely underrepresented in widely used benchmarking efforts. For example, large-scale, multi-task datasets such as the Medical Segmentation Decathlon do not include uterine tasks, reflecting historical data availability rather than clinical need [10]. More broadly, the scarcity of publicly accessible female pelvic MRI datasets with pixel-level annotations has limited the development of standardized evaluation frameworks. The recently released Uterine Myoma MRI Dataset (UMD), comprising 300 annotated sagittal T2-weighted MRI cases with multi-structure labels, represents an important step toward open benchmarking, but also underscores the relative immaturity of this domain [11]. As a result, the uterus remains both clinically relevant and methodologically under-characterized in contemporary segmentation research.

Existing uterine segmentation studies further highlight this gap. Prior work has demonstrated feasibility for uterus-only and multi-structure segmentation, but study designs vary substantially in cohort size, label definitions, and evaluation methodology, limiting comparability across approaches [12–14]. In endometrial cancer, tumor segmentation methods have evolved toward multi-sequence and multitask frameworks yet are still frequently evaluated on single-center datasets with heterogeneous reporting practices [15–17]. For uterine sarcomas, computational imaging research is dominated by radiomics and machine-learning studies that rely on manual segmentations and limited validation, reflecting the scarcity of robust, annotated datasets for rare malignancies [18]. Collectively, this fragmentation constrains not only reproducibility but also our understanding of how segmentation models generalize across uterine imaging contexts.

From a methodological perspective, the medical imaging field has converged on a small number of high-credibility segmentation paradigms. U–Net remains the canonical convolutional encoder–decoder architecture for biomedical image segmentation [19]. nnU–Net demonstrated that much of segmentation performance derives from principled pipeline configuration, encompassing preprocessing, network design, training, and post-processing, automatically adapted to each dataset, thereby establishing a strong and widely adopted baseline [20]. More recently, transformer-based architectures such as Swin–UNETR incorporate hierarchical self-attention to capture long-range spatial dependencies and are increasingly used as high-capacity baselines for 3D medical image segmentation [21, 22]. MedNeXt provides an additional modern convolutional comparison point. It adapts ConvNeXt-style design principles to 3D medical image segmentation using residual inverted bottlenecks and compound scaling to expand effective receptive fields while retaining convolutional locality [23]. However, a robust comparison of these approaches requires not only consistent implementation but also careful evaluation, as overlap-based metrics alone may obscure clinically relevant boundary errors, while segmentation rankings can be sensitive to metric choice, test set composition, and statistical uncertainty [24–26]. Recent work emphasizes the importance of reporting confidence intervals and performing paired statistical comparisons to ensure that observed performance differences are not driven by sampling variability [27].

We therefore designed a controlled multi-dataset evaluation of uterine MRI segmentation spanning benign anatomy, epithelial malignancy, and heterogeneous uterine masses. Three manually configured model families—U-Net, Swin-UNETR, and MedNeXt—were compared under a shared nnU-Net-derived input geometry, while the official nnU-Net framework was retained as a dataset-adaptive system-level reference. The evaluation included one public multiclass dataset, two institutional binary-segmentation cohorts, and a small external cohort relabeled to the public annotation ontology. This design emphasizes comparative robustness rather than proposing a new network architecture.

The study addresses three questions relevant to biomedical image processing: whether model rankings remain stable across anatomically and pathologically distinct segmentation tasks; whether modern convolutional design provides a consistent advantage over a conventional U-Net and a transformer-based baseline; and whether strong internal performance is preserved under cross-institutional and cross-pathology domain shift. The principal contribution is a reproducible, uncertainty-aware comparison across heterogeneous uterine MRI settings using fixed partitions, native-space evaluation, multiple complementary metrics, and paired statistical testing. Figure 1 summarizes the datasets, model families, and evaluation workflow.

**Figure 1.**
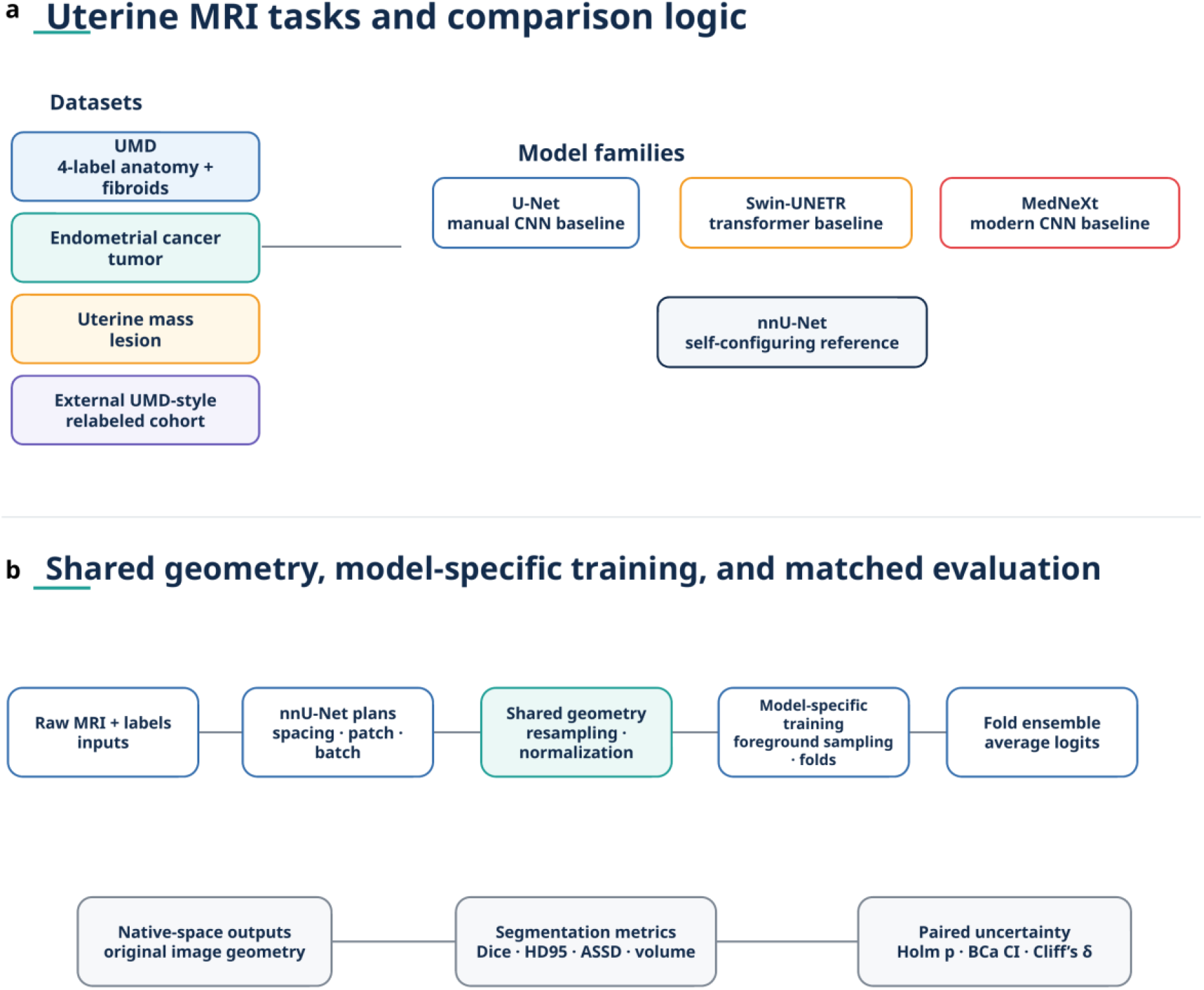
Overview of study design, shared preprocessing geometry, model training, and evaluation. (a) Uterine MRI datasets and segmentation tasks included in the evaluation comprised internal UMD multi-structure segmentation, endometrial cancer tumor segmentation, uterine mass lesion segmentation, and an external UMD-style cohort for out-of-domain testing. (b) U-Net, Swin-UNETR, and MedNeXt were evaluated as manually controlled architectures using shared nnU-Net-derived preprocessing geometry and identical fold assignments, with nnU-Net retained as the dataset-adaptive reference. Model-specific training was followed by fold ensembling, native-space export, overlap-, boundary-, and volume-based evaluation, and paired statistical comparison using bootstrap confidence intervals, Holm-adjusted p values, and effect-size estimation.

## 2. Materials and datasets

### 2.1 Study overview

This study performed a controlled comparative evaluation of segmentation model robustness across three uterine MRI datasets representing distinct anatomical and pathological scenarios, including one public dataset and two institutional cohorts: a multiclass benign uterine anatomy dataset focused on fibroid disease (UMD) [11], an endometrial cancer cohort emphasizing malignant uterine tissue characterization, and a uterine mass cohort designed to capture diagnostically challenging uterine lesions. The uterine mass dataset comprised approximately one-third uterine leiomyosarcoma, one-third atypical fibroids, and one-third typical fibroids, thereby reflecting a clinically relevant mixture of malignant and benign uterine masses. The UMD dataset is publicly available, whereas the endometrial cancer and uterine mass cohorts are institutional datasets that cannot be publicly released because of patient privacy and data-governance restrictions. Accordingly, this work is framed as a controlled comparative evaluation rather than a public leaderboard, with public reproducibility centered on UMD and transparent reporting for the institutional cohorts.

Together, these datasets were selected to capture substantial variability in uterine morphology, lesion burden, class imbalance, and boundary ambiguity, enabling cross-task assessment of model generalizability under clinically heterogeneous conditions. Specifically, the datasets spanned complementary segmentation challenges: (1) multiclass anatomical segmentation with coexisting benign lesions, (2) malignancy-associated distortion of endometrial and myometrial interfaces, and (3) rare lesion segmentation under severe sample limitations.

All datasets were processed independently but evaluated under a unified experimental framework to enable direct comparison of segmentation architectures across tasks.

The institutional components of this retrospective study were conducted under McGill University Health Centre Research Ethics Board approval. Institutional imaging and annotations were handled in accordance with the approved protocol and applicable local data-governance requirements.

### 2.2 UMD dataset: multiclass uterine anatomy and fibroid segmentation

The publicly released UMD dataset consists of pelvic MRI examinations acquired for uterine fibroid assessment and treatment planning [11]. Images include subjects with variable fibroid burden, heterogeneous uterine size, and substantial distortion of normal uterine anatomy, producing a challenging multiclass segmentation setting. Manual annotations were generated for multiple anatomical and lesion classes, including uterine muscular wall, uterine cavity, fibroids and Nabothian cysts. These labels were selected to represent both dominant anatomical structures and smaller clinically relevant lesions, creating marked inter-class differences in spatial extent and prevalence. MRI acquisition was performed using T2-weighted imaging as the principal segmentation sequence, with voxel spacing and field-of-view varying across cases according to clinical protocol.

The UMD dataset includes 300 patients with uterine leiomyoma who underwent pelvic MRI between January 2015 and January 2023. Imaging was acquired on a Philips Ingenia 3.0 T MRI system equipped with a 16-channel coil. The published sagittal T2-weighted protocol reports a slice thickness of 4.0 mm with a 0.4 mm inter-slice gap. Annotation was performed in ITK-SNAP by a team of 8 clinicians with more than 6 years of experience and 3 radiologists with more than 10 years of experience. Each case was labeled independently by two doctors, followed by review after consensus, with additional sequences consulted when lesion identity was uncertain. The study was approved by the scientific research ethics committee of Beijing Shijitan Hospital, Capital Medical University, with exemption of informed consent for the dataset release [11].

The dataset presents several segmentation challenges relevant to model comparison: marked uterine shape variability, large lesion-induced deformation, severe imbalance between dominant and small-volume labels, and occasional near-complete absence of minority structures in some subjects.

### 2.3 Endometrial cancer dataset: tumor segmentation

The institutional endometrial cancer dataset comprises pelvic MRI studies acquired in patients with histologically confirmed endometrial carcinoma. Compared with benign fibroid imaging, this cohort introduces malignant infiltration, irregular endometrial thickening, and partial disruption of normal uterine compartment boundaries.

This dataset was selected because epithelial malignant disease alters both local tissue contrast and global uterine geometry, creating a segmentation problem distinct from benign uterine anatomy. The segmentation objective was binary delineation of lesion versus background; MRI inputs were axial or sagittal T2-weighted images according to the source-hospital protocol.

The endometrial cancer dataset comprised 206 patients with postoperative histopathologic confirmation of endometrial carcinoma from two hospitals. All patients underwent preoperative MRI on 3.0 T GE HealthCare systems, specifically a 3T MR750 and a 3T SIGNA Architect. The segmentation target was binary tumor delineation on T2-weighted MRI. Tumor volumes of interest were manually contoured in ITK-SNAP by a radiologist with 10 years of gynecological oncology MRI experience for all cases. The annotator was aware of the diagnosis of endometrial carcinoma but blinded to histopathologic outcomes.

A key challenge in this cohort is that tumor boundaries may partially overlap normal endometrial interfaces, producing reduced edge contrast and irregular lesion topology.

### 2.4 Uterine mass dataset: lesion segmentation

The privately curated uterine mass dataset comprised multi-center pelvic MRI examinations of patients with uterine masses, including both histopathologically confirmed uLMS and LMs, creating a rare, heterogeneous lesion-segmentation cohort. This setting reflects the clinical challenge of differentiating malignant from benign uterine masses because lesion signal intensity, necrosis, and morphology can overlap substantially across diagnoses.

The uterine mass dataset was derived from a multi-site uterine mass MRI cohort assembled across six institutions and intentionally composed of approximately one-third uLMS, one-third atypical fibroids, and one-third typical fibroids. For the present evaluation, we used a T2-weighted binary lesion-segmentation subset prepared in axial orientation. Site inventory records showed substantial acquisition heterogeneity across the source cohort, including scanners from GE, Philips, and Siemens operating at both 1.5 T and 3 T. T2-weighted imaging was acquired as 2D MRI across sites, with recorded slice thickness values ranging from 3.0 to 7.0 mm and variable inter-slice gap. In the prepared analysis subset, the median voxel spacing was 5.0 × 0.625 × 0.625 mm. These characteristics make the uterine mass cohort particularly valuable for testing segmentation robustness under heterogeneous acquisition conditions and a clinically mixed uterine mass setting.

The final dataset included 234 patients, comprising histopathologically confirmed uLMS cases (n = 77) and LM cases (n = 157), the latter including both typical and atypical imaging presentations. LM cases were confirmed either by histopathology or by imaging stability over a minimum follow-up period of two years. For the primary uterine mass evaluation, all cases were represented using a common binary lesion-segmentation target so that the task focused on lesion delineation rather than diagnosis.

For the purposes of this study, the segmentation task was formulated as binary lesion delineation (lesion versus background). Model inputs consisted of T2-weighted MRI, selected for consistency across institutions and for its central role in uterine imaging. Ground-truth annotations were generated by a radiologist with 10 years of gynecological oncology MRI experience for all cases using a standardized protocol applied across sites.

The dataset presents several challenges relevant to model evaluation, including limited sample size, marked lesion heterogeneity, necrotic and cystic components, and variability in acquisition protocols across institutions. These characteristics make the uterine mass cohort particularly well suited for assessing segmentation performance under small-data conditions and for evaluating the robustness of model architectures to domain shift.

### 2.5 External UMD-style test cohort for cross-institution evaluation

This cohort consisted of 12 additional sagittal cases drawn from the uterine mass dataset and excluded from the primary uterine mass analyses. These cases were selected specifically because their acquisition plane matched the sagittal UMD protocol. Selection was based on imaging plane rather than lesion subtype, size, or other matching criteria, since the purpose of this experiment was not to create a lesion-matched comparison set, but to evaluate transfer of UMD-trained models under a deliberately challenging anatomical and pathological domain shift. All UMD-style relabeling was performed by the same radiologist with 10 years of gynecological oncology MRI experience who generated the institutional cohort segmentations, using label descriptions matched as closely as possible to the published UMD annotation protocol. Pathology labels for the 12 relabeled external cases were not available for this analysis. The cases were acquired on mixed scanner platforms and protocols; available metadata indicated multi-vendor, multi-field-strength 2D T2-weighted imaging, with resolution and slice parameters differing from UMD, but scanner-specific effects could not be separated because of the small sample size and mixed provenance. Available characteristics are summarized in Table 1 where available.

**Table 1.** Imaging acquisition, segmentation plane, and annotation characteristics across datasets.

| Dataset | Clinical task | MRI sequence | Segmentation plane | Scanner / vendor | Field strength | Coil | Key acquisition details | Annotation target(s) |
| --- | --- | --- | --- | --- | --- | --- | --- | --- |
| UMD | Benign multi-structure uterine anatomy and fibroid segmentation | T2-weighted MRI | Sagittal | Philips Ingenia | 3.0 T | 16-channel coil | TR 4200 ms; TE 130 ms; flip angle 90°; voxel size $0.8 \times 0.8 \times 4.0$ mm; slice gap 0.4 mm; field of view $24 \times 24$ cm | Uterine muscular wall, uterine cavity, myomas, Nabothian cysts |
| Endometrial cancer | Endometrial cancer tumor segmentation | T2-weighted MRI | Axial (Hospital A) and sagittal (Hospital B) | 3T MR750 (Hospital A) and 3T SIGNA Architect (Hospital B), GE HealthCare | 3.0 T | 32-channel Body Array coil (Hospital A); 30-channel Adaptive Imaging Receive coil (Hospital B) | Median voxel spacing $4.0 \times 0.4297 \times 0.4297$ mm | Tumor |
| Uterine mass | Uterine mass / lesion segmentation | T2-weighted MRI | Axial | Multi-site cohort including GE, Philips, and Siemens systems | 1.5 T and 3.0 T | Site-dependent | Multi-site 2D T2-weighted MRI; T2 slice thickness ranged from 3.0 to 7.0 mm across contributing sites; variable slice gap; analysis subset median voxel spacing $5.0 \times 0.625 \times 0.625$ mm | Lesion |

### 2.6 Dataset heterogeneity as an experimental design principle

Rather than treating inter-dataset heterogeneity as a limitation, this study explicitly uses heterogeneity as an experimental design feature. This design allows evaluation of whether segmentation model ranking remains stable across uterine tasks or changes according to anatomical complexity, lesion rarity, and pathology-induced deformation. The main characteristics of the included datasets and segmentation tasks are summarized in Table 2.

**Table 2.** Dataset characteristics.

| Dataset | Clinical task | n | MRI sequence | Labels | Pathology type | Annotation target |
| --- | --- | --- | --- | --- | --- | --- |
| UMD | Benign multi-structure uterus | 300 | T2-weighted MRI | 4 | Fibroids / benign anatomy | wall, cavity, myoma, Nabothian cyst |
| Endometrial cancer | Endometrial carcinoma | 206 | T2-weighted MRI | 1 | epithelial malignancy | tumor |
| Uterine mass | Uterine mass segmentation | 234 | T2-weighted MRI | 1 | leiomyosarcoma, atypical fibroids, typical fibroids | lesion |

## 3. Methods

### 3.1 Experimental design

Three manually controlled architectures and one dataset-adaptive reference framework were evaluated across all datasets: a manually configured MONAI U-Net pipeline [28], the self-configuring nnU-Net reference framework [20], a MONAI Swin-UNETR transformer-based architecture [28], and the MedNeXt architecture implemented in MONAI [23]. These models were selected to represent two complementary levels of comparison, first, manually configured architecture-level differences among U-Net, Swin-UNETR, and MedNeXt, and second, system-level performance against a fully self-configuring reference pipeline. All models were evaluated under identical dataset partitions for each task; architecture-dependent differences were interpreted primarily among the three manually controlled architectures, while nnU-Net was treated as the dataset-adaptive reference. For each dataset, training and inference were performed independently using task-specific labels while preserving a common evaluation framework. The fourth model, MedNeXt, was included to test whether a modern ConvNeXt-inspired 3D convolutional architecture could improve over the conventional U-Net baseline while retaining convolutional inductive bias [23].

### 3.2 Common preprocessing framework

To ensure comparability across models, all experiments were anchored to a common nnU-Net-derived preprocessing geometry. No additional N4 bias-field correction or manual intensity preprocessing was applied. For the MONAI-based models (U-Net, Swin-UNETR, and MedNeXt) [28], input images were converted using nnU-Net preprocessing plans generated from the corresponding dataset configuration. Target spacing and the nominal patch and batch configurations were derived from the dataset-specific nnU-Net plans file, with architecture-specific spatial-divisibility adjustments applied where required by Swin-UNETR and MedNeXt. All models otherwise operated on the same resampled voxel geometry. This design was intended to prevent model comparisons from being confounded by different resampling, cropping, or intensity-normalization conventions.

Importantly, this shared preprocessing does not mean that the MONAI U-Net was a replication of the nnU-Net method. The U-Net baseline used the same input geometry as nnU-Net, but it did not use nnU-Net’s full self-configuring pipeline, automatically selected network topology, deep-supervision strategy, augmentation configuration, optimizer schedule, or end-to-end dataset-specific training recipe. It should therefore be interpreted as a controlled convolutional baseline trained under matched preprocessing conditions, rather than as an independently reimplemented nnU-Net. In contrast, the nnU-Net reference model used the official nnU-Net framework, including its dataset-adaptive configuration of preprocessing, architecture, training, and inference procedures.

Intensity normalization followed the default nnU-Net preprocessing logic, with per-case z-score normalization applied to the single T2-weighted MRI channel. For MONAI models, training data were loaded from nnU-Net preprocessed arrays while preserving ignore-label voxels where applicable. Consequently, the MONAI model comparisons should be interpreted as comparisons of architecture and training configuration under a shared nnU-Net-derived input representation, rather than as comparisons of fully independent preprocessing pipelines.

Training, inference, validation, and reporting code is publicly available at https://github.com/AIPHL-McGill/UterusSegBenchmark. The repository provides the MONAI training scripts, five-fold launcher, nnU-Net v2.6.2-compatible preprocessing and export workflow, benchmark validator, environment specification, and task-specific command examples. The environment specifies Python 3.10, PyTorch 2.0 or later, MONAI 1.5.0 or later, SimpleITK 2.3 or later, and nnU-Net v2. Dataset-specific plans and protected split files are not redistributed, but the executable pipeline and configuration interface used in this study are available.

### 3.3 Cross-validation and data partitioning

Five-fold cross-validation was performed within each development set using predefined fold assignments generated before training. Fold membership was held constant across all architectures, and the fixed held-out test sets were excluded from model fitting, checkpoint selection, and hyperparameter decisions. The development/test partitions were 270/30 cases for UMD, 163/43 for endometrial cancer, and 193/41 for the uterine mass dataset. The external UMD-style cohort comprised 12 test-only cases. The same split files were provided to all MONAI-based models and to nnU-Net to preserve strict comparability.

### 3.4 U-Net implementation

The convolutional baseline used a 3D U-Net implementation in MONAI. To account for anisotropic pelvic MRI voxel spacing, the network was trained on nnU-Net-matched patch geometry and preprocessing plans. The architecture used five encoder-decoder stages with channel progression (32, 64, 128, 256, 512), strides (2, 2, 2, 2), two residual units per stage, leaky-ReLU activations, instance normalization, and no dropout. The network operated on single-channel volumetric T2-weighted MRI input and produced either binary or multiclass output depending on the dataset label structure. Training used stochastic gradient descent with momentum = 0.99, Nesterov acceleration, weight decay = 3 × 10⁻⁵, and polynomial learning-rate decay (power = 0.9), with an initial learning rate of 10⁻².

### 3.5 Swin-UNETR implementation

The transformer-based model used Swin-UNETR implemented in MONAI. As with the U-Net, the architecture was adapted to nnU-Net-derived patch geometry, while enforcing spatial divisibility constraints required by the Swin hierarchy. The default configuration used feature size = 12, attention heads = [3, 6, 12, 24], depths = [2, 2, 2, 1], and window size = 7. For anisotropic datasets, the training code applied spacing-aware adjustments to the region of interest and, when supported by the installed MONAI version, anisotropy-aware patch and window settings. Unlike the convolutional baseline, Swin-UNETR was optimized using AdamW with learning rate = 2 × 10⁻⁴, β = (0.9, 0.999), and weight decay = 5 × 10⁻². This optimizer change was introduced because preliminary experiments showed that the nnU-Net-style SGD schedule produced unstable convergence for transformer training.

### 3.6 MedNeXt implementation

MedNeXt was implemented in MONAI as a modernized convolutional segmentation model. Unlike Swin-UNETR, MedNeXt does not use self-attention; instead, it increases contextual modeling capacity through ConvNeXt-style 3D convolutional blocks, residual inverted bottlenecks, and scalable receptive-field design. This architecture was included to test whether transformer-era convolutional design principles provide advantages for anisotropic, modest-sized uterine MRI datasets while preserving the locality and data efficiency of convolutional models. The default experiment used the MedNeXt-S configuration with initial filters = 32, kernel size = 3, group normalization, residual connections enabled, and no global response normalization. MedNeXt used the same nnU-Net-derived preprocessing geometry, foreground sampling, loss formulation, validation protocol, and fold ensembling strategy as the other MONAI-based models. As a convolutional model, it was optimized with the same stochastic-gradient-descent schedule used for the U-Net baseline.

### 3.7 Patch sampling and foreground balancing

A key methodological challenge across uterine datasets was severe label imbalance, particularly for small lesion structures and rare classes. To address this, MONAI-based training used an nnU-Net-inspired foreground oversampling strategy implemented through a custom patch sampler. Effective foreground probability was quantized in nnU-Net-like fashion so that, for batch size = 2, exactly one patch per batch was forced to contain foreground. Forced-foreground patches used class-aware sampling, with preferential sampling of rarer labels through cached foreground coordinates. This strategy was especially important for UMD, where minority labels such as Nabothian cysts occupied very small spatial fractions.

### 3.8 Loss formulation

Training used a composite loss combining ignore-aware soft Dice loss and cross-entropy. For multiclass segmentation, the Dice component excluded background to mirror standard nnU-Net conventions, while the cross-entropy term was computed with the specified ignore index. For ignored labels, masked Dice statistics were computed so that excluded voxels did not contribute to gradient updates.

### 3.9 Validation protocol

Validation was performed using full-case inference rather than patch-level validation. Every 50 epochs, full-volume sliding-window inference was applied to up to eight validation cases selected from the fold-specific validation split. Validation Dice was computed per class and aggregated into mean foreground Dice using nnU-Net-style foreground averaging. The best checkpoint per fold was selected according to mean foreground validation Dice.

### 3.10 nnU-Net reference model

The nnU-Net reference used the official nnU-Net full-resolution 3D configuration (v2.6.2). For each dataset, nnU-Net automatically determined target spacing, patch size, batch size, normalization policy, and deep-supervision strategy. This framework served as an automated reference because it jointly optimizes preprocessing and architecture rather than requiring manual tuning.

### 3.11 Inference and fold ensembling

For MONAI-based models, inference was performed using sliding-window prediction with nnU-Net-compatible export geometry. All fold checkpoints were ensembled by averaging logits before final label assignment. Sliding-window inference used a batch size of 1, Gaussian blending, and overlap of 0.5. Predictions were exported in the original image geometry using nnU-Net v2.6.2 preprocessing and export routines to preserve voxel alignment with the reference annotations. This ensured that MONAI outputs and nnU-Net outputs were directly comparable at the voxel level.

### 3.12 Evaluation metrics and statistical comparison

Performance was evaluated independently for each dataset using Dice similarity coefficient, 95th-percentile Hausdorff distance (HD95), average symmetric surface distance (ASSD), and absolute volume difference. HD95 was calculated in physical millimeters as the maximum of the two directed 95th-percentile surface-to-surface distances. HD95 and ASSD were calculated only when both the reference and predicted masks were nonempty; reference-present cases with empty predictions were recorded as complete detection failures and excluded from the conditional distance summaries rather than assigned an arbitrary penalty. Final test-set evaluation was performed on fixed held-out test sets for each dataset, separate from the fold-specific validation cases used for checkpoint selection. For multiclass tasks, metrics were computed per label and summarized using macro-averaged subject-level means. Pairwise model comparisons used paired bootstrap resampling (5,000 iterations) with two-sided bootstrap *p*-values and bias-corrected and accelerated (BCa) confidence intervals. To control multiplicity, bootstrap *p*-values were Holm-adjusted within each dataset across the six pairwise model comparisons. Effect size was quantified using Cliff’s delta. This paired design ensured that model differences were assessed on identical cases rather than aggregated group summaries.

### 3.13 External evaluation protocol

Beyond the primary within-dataset evaluation, we performed an additional external evaluation to assess generalization of the UMD-trained models. For this analysis, models trained on UMD were applied without retraining or adaptation to the relabeled external UMD-style cohort described in Section 2 and drawn from the uterine mass cohort. The cohort matched UMD in image orientation and annotation ontology but differed in institutional source, scanner characteristics, voxel spacing, pathology, and case mix. It therefore provided a targeted test of cross-institution robustness while holding orientation and label definitions approximately constant, rather than isolating a single source of domain shift. Performance was quantified using the same evaluation framework as the primary evaluation, including Dice similarity coefficient, HD95, ASSD, and volumetric disagreement. Detailed acquisition characteristics are reported in Table 1 when available.

## 4. Results

### 4.1 Architecture-level performance and dataset-adaptive reference

Results were interpreted in two complementary ways. First, as an architecture-level comparison among the manually controlled U-Net, Swin-UNETR, and MedNeXt models under matched preprocessing; and second, as a system-level comparison against the self-configuring nnU-Net reference. Within the manually controlled architecture comparison, MedNeXt consistently performed best by macro-Dice. Against this architecture-level comparison, nnU-Net achieved the strongest overall segmentation performance in every dataset, including the external UMD-style validation cohort. Overall quantitative performance across datasets and models is summarized in Table 3. The magnitude of the system-level nnU-Net advantage varied by task, with the clearest separation from MedNeXt observed in the multi-structure UMD setting and smaller differences in the binary segmentation tasks. Pairwise statistical comparisons of macro-Dice between models are reported in Table 4.

**Table 3.** Overall macro-averaged segmentation performance for each model across the three primary uterine MRI datasets and the external UMD-style validation cohort.

| Dataset | Model | n | Dice<br>(mean ± SD) | HD95 (mm)<br>(mean ± SD) | ASSD (mm)<br>(mean ± SD) | Absolute volume<br>difference (mL)<br>(mean ± SD) |
| --- | --- | --- | --- | --- | --- | --- |
| UMD | U-Net | 30 | 0.647 ± 0.135 | 13.34 ± 10.16 | 4.14 ± 3.36 | 7.33 ± 7.08 |
| UMD | Swin-UNETR | 30 | 0.592 ± 0.140 | 15.83 ± 11.04 | 5.03 ± 4.17 | 9.00 ± 9.28 |
| UMD | nnU-Net | 30 | **0.761 ± 0.114** | 10.25 ± 10.68 | 3.03 ± 2.60 | 3.66 ± 3.62 |
| UMD | MedNeXt | 30 | 0.722 ± 0.114 | 10.62 ± 11.14 | 3.22 ± 2.75 | 5.47 ± 6.31 |
| Endometrial cancer | U-Net | 43 | 0.689 ± 0.241 | 20.09 ± 32.58 | 6.14 ± 11.68 | 4.30 ± 6.41 |
| Endometrial cancer | Swin-UNETR | 43 | 0.632 ± 0.292 | 14.49 ± 28.99 | 3.64 ± 5.37 | 5.93 ± 9.24 |
| Endometrial cancer | nnU-Net | 43 | **0.746 ± 0.257** | 10.24 ± 16.21 | 2.95 ± 3.81 | 2.60 ± 3.59 |
| Endometrial cancer | MedNeXt | 43 | 0.726 ± 0.256 | 15.54 ± 31.88 | 4.64 ± 9.22 | 3.61 ± 5.84 |
| Uterine mass | U-Net | 41 | 0.744 ± 0.262 | 46.19 ± 53.62 | 9.24 ± 10.35 | 181.63 ± 280.61 |
| Uterine mass | Swin-UNETR | 41 | 0.733 ± 0.286 | 43.88 ± 46.23 | 10.02 ± 10.76 | 190.88 ± 279.06 |
| Uterine mass | nnU-Net | 41 | **0.814 ± 0.268** | 25.48 ± 38.43 | 6.27 ± 9.12 | 95.91 ± 136.69 |
| Uterine mass | MedNeXt | 41 | 0.789 ± 0.242 | 29.68 ± 38.71 | 6.08 ± 5.90 | 121.46 ± 163.83 |
| External UMD-style cohort | U-Net | 12 | 0.396 ± 0.225 | 52.71 ± 32.38 | 13.39 ± 9.23 | 413.71 ± 439.24 |
| External UMD-style cohort | Swin-UNETR | 12 | 0.287 ± 0.225 | 71.12 ± 53.96 | 22.29 ± 17.84 | 423.17 ± 448.57 |
| External UMD-style cohort | nnU-Net | 12 | **0.542 ± 0.212** | 38.12 ± 29.68 | 9.29 ± 7.31 | 291.80 ± 340.76 |
| External UMD-style cohort | MedNeXt | 12 | 0.490 ± 0.230 | 47.67 ± 35.74 | 10.77 ± 8.81 | 357.00 ± 393.53 |

Note: HD95 and ASSD are conditional on nonempty reference and prediction masks. Finite HD95 observations (U-Net/Swin-UNETR/nnU-Net/MedNeXt) were 100/99/100/101 of 101 label-present observations in UMD; 39/38/38/37 of 44 in the external UMD-style cohort; 43/41/40/42 of 43 in endometrial cancer; and 41/41/39/40 of 41 in the uterine mass dataset. The number of complete detection failures equals the label-present denominator minus the finite observation count for each model. Complete misses were excluded from the distance summaries rather than assigned an arbitrary penalty.

**Table 4.** Pairwise statistical comparisons of macro-Dice across models within each dataset.

| Dataset | Comparison | Mean Dice difference | BCa 95% CI | Holm-adjusted p-value | Cliff's $\delta$ |
| --- | --- | --- | --- | --- | --- |
| UMD | U-Net vs Swin-UNETR | 0.055 | [0.028, 0.091] | <0.006 | 0.222 |
| UMD | U-Net vs nnU-Net | -0.113 | [-0.150, -0.085] | <0.006 | -0.482 |
| UMD | Swin-UNETR vs nnU-Net | -0.169 | [-0.205, -0.136] | <0.006 | -0.640 |
| UMD | U-Net vs MedNeXt | -0.075 | [-0.097, -0.057] | <0.006 | -0.316 |
| UMD | Swin-UNETR vs MedNeXt | -0.130 | [-0.165, -0.101] | <0.006 | -0.507 |
| UMD | nnU-Net vs MedNeXt | 0.039 | [0.016, 0.063] | 0.001 | 0.209 |
| Endometrial cancer | U-Net vs Swin-UNETR | 0.058 | [0.003, 0.123] | 0.092 | 0.091 |
| Endometrial cancer | U-Net vs nnU-Net | -0.057 | [-0.086, -0.013] | 0.024 | -0.286 |
| Endometrial cancer | Swin-UNETR vs nnU-Net | -0.114 | [-0.192, -0.063] | <0.006 | -0.346 |
| Endometrial cancer | U-Net vs MedNeXt | -0.037 | [-0.062, -0.007] | 0.039 | -0.187 |
| Endometrial cancer | Swin-UNETR vs MedNeXt | -0.095 | [-0.170, -0.043] | <0.006 | -0.243 |
| Endometrial cancer | nnU-Net vs MedNeXt | 0.020 | [-0.004, 0.039] | 0.092 | 0.118 |
| Uterine mass | U-Net vs Swin-UNETR | 0.011 | [-0.041, 0.081] | 0.715 | -0.057 |
| Uterine mass | U-Net vs nnU-Net | -0.069 | [-0.121, -0.001] | 0.120 | -0.394 |
| Uterine mass | Swin-UNETR vs nnU-Net | -0.081 | [-0.147, -0.047] | <0.006 | -0.354 |
| Uterine mass | U-Net vs MedNeXt | -0.045 | [-0.094, -0.003] | 0.135 | -0.185 |
| Uterine mass | Swin-UNETR vs MedNeXt | -0.056 | [-0.103, -0.028] | <0.006 | -0.142 |
| Uterine mass | nnU-Net vs MedNeXt | 0.025 | [-0.008, 0.056] | 0.248 | 0.286 |
| External UMD-style cohort | U-Net vs Swin-UNETR | 0.110 | [0.045, 0.238] | <0.006 | 0.333 |
| External UMD-style cohort | U-Net vs nnU-Net | -0.145 | [-0.235, -0.091] | <0.006 | -0.431 |
| External UMD-style cohort | Swin-UNETR vs nnU-Net | -0.255 | [-0.390, -0.154] | <0.006 | -0.625 |
| External UMD-style cohort | U-Net vs MedNeXt | -0.094 | [-0.164, -0.056] | <0.006 | -0.368 |
| External UMD-style cohort | Swin-UNETR vs MedNeXt | -0.203 | [-0.311, -0.116] | <0.006 | -0.514 |
| External UMD-style cohort | nnU-Net vs MedNeXt | 0.052 | [-0.014, 0.087] | 0.044 | 0.139 |

Note: $p$ -values are two-sided paired-bootstrap $p$ -values adjusted within each dataset using Holm correction across the six pairwise model comparisons. BCa intervals are unadjusted 95% paired-bootstrap intervals.

In UMD, MedNeXt was the strongest manually controlled architecture (macro-Dice 0.722), outperforming U-Net (0.647) and Swin-UNETR (0.592), while the dataset-adaptive nnU-Net reference achieved the highest macro-Dice (0.761). nnU-Net also achieved the lowest conditional mean HD95 (10.25 mm) and ASSD (3.03 mm). MedNeXt ranked second for overlap and ASSD, with a similar conditional mean HD95 of 10.62 mm and ASSD of 3.22 mm. Its absolute volume disagreement (5.47 mL) remained higher than nnU-Net (3.66 mL) and lower than U-Net (7.33 mL) and Swin-UNETR (9.00 mL).

In the endometrial cancer dataset, MedNeXt again led the manually controlled architectures and narrowed the gap to the nnU-Net reference. nnU-Net achieved the highest macro-Dice (0.746), followed by MedNeXt (0.726), U-Net (0.689), and Swin-UNETR (0.632). After Holm correction, MedNeXt remained higher than U-Net (mean Dice difference 0.037, BCa 95% CI 0.007 to 0.062, adjusted p = 0.039 when expressed as MedNeXt minus U-Net) and Swin-UNETR (0.095, BCa 95% CI 0.043 to 0.170, adjusted p < 0.006). The nnU-Net-MedNeXt difference was smaller and not statistically robust by BCa confidence interval or adjusted *p*-value (0.020, BCa 95% CI -0.004 to 0.039, adjusted p = 0.092). Thus, the endometrial cancer dataset showed a compact ordering in which the strongest manually controlled architecture approached nnU-Net overlap performance while retaining higher boundary error.

In the uterine mass dataset, MedNeXt was also the strongest manually controlled architecture (macro-Dice 0.789), followed by U-Net (0.744) and Swin-UNETR (0.733), while nnU-Net yielded the highest macro-Dice (0.814). nnU-Net achieved the lowest conditional mean HD95 (25.48 mm) and absolute volume difference (95.91 mL), whereas MedNeXt achieved the lowest mean ASSD (6.08 mm versus 6.27 mm for nnU-Net) and the second-lowest HD95 (29.68 mm). Taken together, these findings indicate that MedNeXt was the strongest manually controlled architecture, whereas the self-configuring nnU-Net pipeline remained the most robust system-level reference across benign multi-label anatomy, epithelial malignancy, and heterogeneous uterine masses. Representative qualitative examples from the internal and external evaluation tasks are shown in Figure 2.

**Figure 2.**
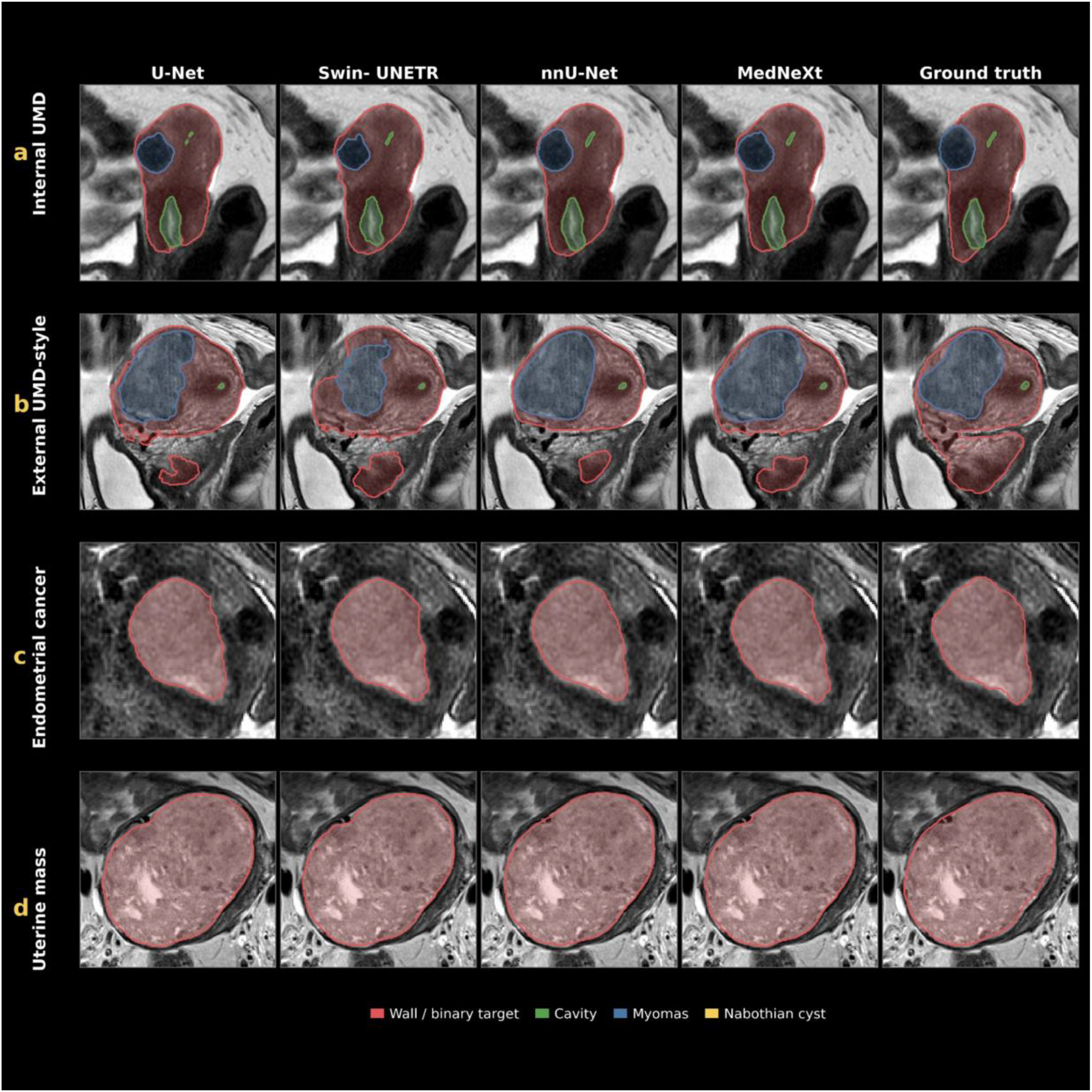
Representative qualitative segmentation outputs across the three primary uterine MRI tasks and the external evaluation. (a) Internal UMD multi-structure segmentation. (b) External UMD-style cohort relabeled to the UMD protocol. (c) Endometrial cancer tumor segmentation. (d) Uterine mass lesion segmentation. For each panel, representative T2-weighted MRI slices are shown together with predictions from U-Net, Swin-UNETR, nnU-Net, and MedNeXt, alongside expert reference annotations.

### 4.2 Label-specific performance in UMD

The multiclass UMD dataset showed marked label dependence in model performance. For uterine muscular wall, Dice increased from 0.805 with U-Net and 0.769 with Swin-UNETR to 0.875 with nnU-Net, with MedNeXt ranking second at 0.848. For uterine cavity, the corresponding Dice values were 0.695, 0.674, 0.782, and 0.753. The largest improvement occurred for leiomyomas, where nnU-Net reached 0.669 compared with 0.573 for MedNeXt, 0.464 for U-Net, and 0.396 for Swin-UNETR, consistent with the greater morphological variability of this label. Nabothian cyst remained the most challenging target overall; MedNeXt achieved the highest numerical Dice for this label (0.661), followed by nnU-Net (0.607), U-Net (0.533), and Swin-UNETR (0.359), although only 11 positive cases were available. These results indicate that the performance gap widened as structures became smaller, rarer, or more heterogeneous, and that MedNeXt was particularly competitive for the smallest UMD label. The distribution of Dice scores across datasets, models, and UMD label categories is summarized in Figure 3.

**Figure 3.**
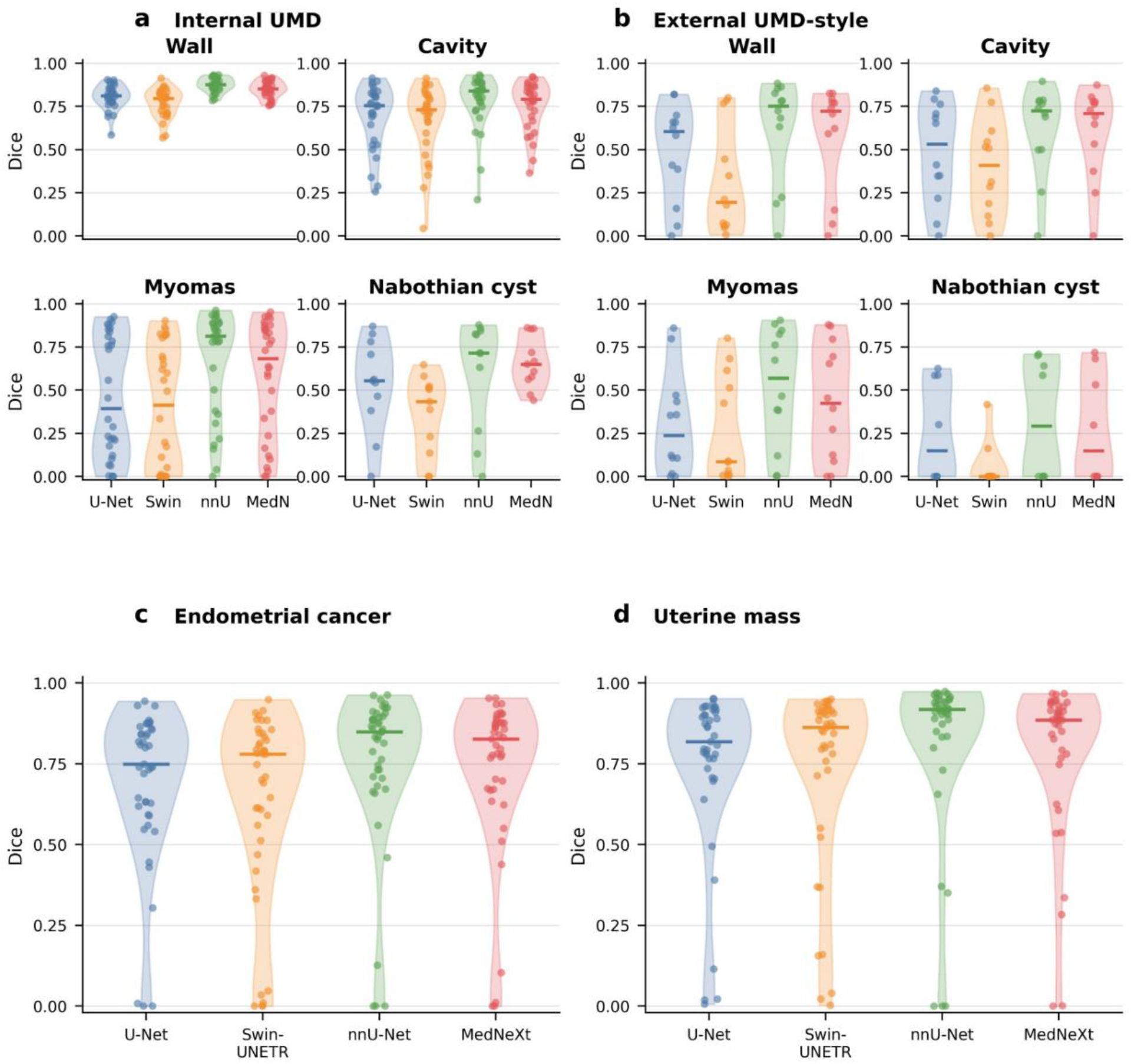
Distribution of Dice similarity coefficients across datasets and model architectures. (a) Internal UMD label-wise Dice distributions for multi-structure segmentation. (b) External UMD-style cohort relabeled to the UMD annotation protocol. (c) Endometrial cancer tumor Dice distributions. (d) Uterine mass lesion Dice distributions. Violin plots summarize case-level Dice similarity coefficients for each model, with central markers indicating mean performance and distribution width reflecting inter-case variability. For label-wise UMD displays, reference-present label observations were summarized where applicable.

### 4.3 Statistical comparison in UMD

Pairwise bootstrap analysis confirmed both levels of the UMD comparison after within-dataset Holm correction. At the system level, nnU-Net exceeded MedNeXt by a smaller but statistically robust margin (mean Dice difference 0.039, BCa 95% CI 0.016 to 0.063, Holm-adjusted *p* = 0.001), whereas at the architecture level MedNeXt exceeded U-Net (0.075, BCa 95% CI 0.057 to 0.097, Holm-adjusted *p* < 0.006 when expressed as MedNeXt minus U-Net) and Swin-UNETR (0.130, BCa 95% CI 0.101 to 0.165, Holm-adjusted *p* < 0.006). At the label level, nnU-Net remained higher than MedNeXt for uterine muscular wall, cavity, and myomas. For Nabothian cyst, MedNeXt was numerically higher than nnU-Net, but this difference was not statistically significant (mean difference 0.053 when expressed as MedNeXt minus nnU-Net, BCa 95% CI crossing zero, *p* = 0.530).

### 4.4 Endometrial cancer dataset performance and statistical comparison

In the endometrial cancer dataset, MedNeXt again led the manually configured architectures and narrowed the gap to the nnU-Net reference. nnU-Net achieved the highest macro-Dice (0.746), followed by MedNeXt (0.726), U-Net (0.689), and Swin-UNETR (0.632). After within-dataset Holm correction, MedNeXt remained higher than U-Net (mean Dice difference 0.037, BCa 95% CI 0.007 to 0.062, adjusted p = 0.039 when expressed as MedNeXt minus U-Net) and Swin-UNETR (0.095, BCa 95% CI 0.043 to 0.170, adjusted p < 0.006). The difference between nnU-Net and MedNeXt was smaller and not statistically robust (0.020, BCa 95% CI -0.004 to 0.039, adjusted p = 0.092). Thus, the strongest manually configured architecture approached nnU-Net overlap performance in this binary task, although nnU-Net retained lower conditional boundary error.

### 4.5 Uterine mass dataset performance and statistical comparison

The uterine mass dataset was the most favorable setting for modern convolutional scaling relative to the other manually controlled architectures, although nnU-Net remained the top-performing reference framework. Mean macro-Dice reached 0.814 for nnU-Net, followed by 0.789 for MedNeXt, 0.744 for U-Net, and 0.733 for Swin-UNETR.

Pairwise analysis showed that MedNeXt was descriptively higher than U-Net (mean Dice difference 0.045, BCa 95% CI 0.003 to 0.094 when expressed as MedNeXt minus U-Net), although this comparison did not survive within-dataset Holm correction (adjusted *p* = 0.135). MedNeXt remained higher than Swin-UNETR after correction (0.056, BCa 95% CI 0.028 to 0.103, adjusted *p* < 0.006). The difference between nnU-Net and MedNeXt was smaller and not statistically robust by BCa confidence interval or adjusted *p*-value (0.025, BCa 95% CI -0.008 to 0.056, adjusted p = 0.248).

This indicates that transformer-based modeling did not confer a consistent advantage for heterogeneous uterine mass segmentation under the present training conditions, whereas the MedNeXt results suggest that transformer-inspired convolutional scaling may be better suited to this modest-sized, anisotropic pelvic MRI setting. Overall, the uterine mass dataset reinforces the broader two-level result, one that MedNeXt provided the strongest architecture-level alternative, and two nnU-Net remained the most robust dataset-adaptive reference.

### 4.6 External UMD-style validation of UMD-trained models

To assess cross-domain generalization while holding image orientation and annotation ontology approximately constant, we evaluated the UMD-trained models on an external cohort of 12 sagittal uterine mass acquisitions relabeled to match the UMD label set.

All models degraded substantially on this external set, but the two-level ranking was preserved. nnU-Net achieved the highest macro-Dice (0.542), followed by MedNeXt (0.490), U-Net (0.396), and Swin-UNETR (0.287). The same ordering was observed for the conditional boundary metrics: nnU-Net had the lowest mean HD95 (38.12 mm) and ASSD (9.29 mm), followed by MedNeXt (HD95 47.67 mm; ASSD 10.77 mm), U-Net (52.71 mm; 13.39 mm), and Swin-UNETR (71.12 mm; 22.29 mm). Absolute volume disagreement remained very large for all models but was lowest for nnU-Net (291.80 mL), followed by MedNeXt (357.00 mL), U-Net (413.71 mL), and Swin-UNETR (423.17 mL).

At the label level, external generalization was moderate for the larger anatomical structures but weaker for pathology-dependent or low-prevalence labels. Pairwise overall comparisons showed that MedNeXt remained higher than U-Net (mean Dice difference 0.094, BCa 95% CI 0.056 to 0.164, Holm-adjusted *p* < 0.006 when expressed as MedNeXt minus U-Net) and Swin-UNETR (0.203, BCa 95% CI 0.116 to 0.311, adjusted *p* < 0.006). The difference between nnU-Net and MedNeXt was smaller and should be interpreted cautiously because the BCa confidence interval crossed zero (0.052, BCa 95% CI -0.014 to 0.087), despite an adjusted bootstrap *p*-value of 0.044. These findings indicate that even when image orientation and label space are matched, uterine MRI segmentation remains highly sensitive to cross-institution and cross-pathology domain shift. Complementary boundary-based comparisons using 95th-percentile Hausdorff distance are shown in Figure 4.

**Figure 4.**
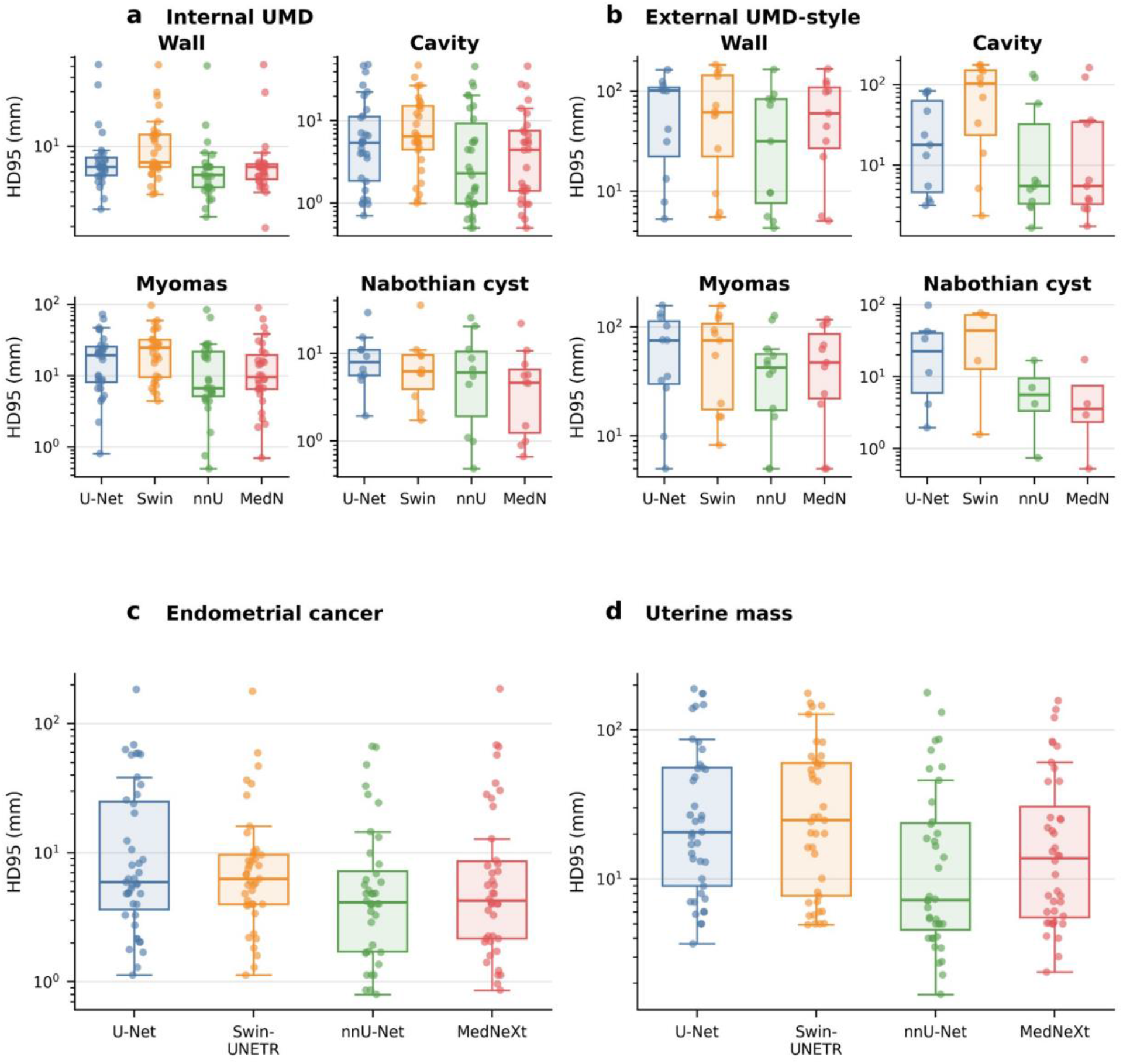
Boundary agreement measured by 95th-percentile Hausdorff distance (HD95). (a) Internal UMD boundary error across anatomical labels. (b) External UMD-style cohort relabeled to the UMD annotation protocol. (c) Endometrial cancer tumor boundary error. (d) Uterine mass lesion boundary error. Boxplots with overlaid case-level values show conditional HD95 in millimeters for each model on a logarithmic scale, where lower values indicate closer boundary agreement with expert annotations. HD95 was calculated only when both reference and prediction masks were nonempty; complete misses were excluded from the displayed distance distributions and are enumerated in the Table 3 note. Compared with overlap-based metrics, HD95 highlights localized contour failures and differences in robustness to irregular lesion margins and small anatomical structures.

## 5. Discussion

This study evaluated the robustness of four established segmentation paradigms across three clinically distinct uterine MRI tasks and a separate external domain-shift cohort. Two findings were consistent. First, MedNeXt was the strongest manually configured architecture, indicating that modern convolutional block design can improve over a conventional U-Net and, under the present training conditions, over Swin-UNETR. Second, nnU-Net remained the strongest complete system across all internal datasets and external testing. The results therefore distinguish an architecture-level conclusion from a pipeline-level conclusion, namely that backbone design mattered, but dataset-adaptive configuration was at least as important as architectural family.

Uterine MRI provides a demanding test of segmentation robustness because target appearance varies across both anatomy and disease. Fibroids can markedly deform uterine compartments, endometrial carcinoma can obscure the endometrial-myometrial interface, and heterogeneous uterine masses can contain necrotic, cystic, and solid components [2,29,30]. These factors interact with anisotropic acquisition, class imbalance, and differences in label ontology. Consequently, performance on one uterine task should not be assumed to transfer to another, even when all tasks use T2-weighted MRI.

The performance of nnU-Net is consistent with its design as a self-configuring segmentation system [20]. Its advantage cannot be attributed to a single component because preprocessing, network topology, augmentation, deep supervision, optimization, and inference are configured jointly. The present comparison therefore shows that the complete adaptive pipeline provided a reliable reference across datasets with different spacing, target size, label structure, and pathology. This distinction is important when manually configured networks are compared with nnU-Net as the comparison is between architectures under a shared input representation on one hand and a full adaptive system on the other.

MedNeXt provided the clearest architecture-level result. Its ConvNeXt-inspired design was descriptively stronger than U-Net and Swin-UNETR across datasets, with statistically robust corrected differences in UMD and endometrial cancer and against Swin-UNETR in the uterine mass cohort. The smaller separation from nnU-Net in the two binary tasks suggests that modern convolutional scaling is a competitive option when the target is a single lesion class. The larger gap in multiclass UMD segmentation, particularly for myomas, indicates that architecture-level improvements did not fully compensate for the demands of simultaneous multi-structure segmentation and severe class imbalance.

The multi-metric evaluation also exposed failure modes that Dice alone would not reveal. Conditional HD95 and ASSD remained large in malignant and external cohorts even when mean Dice was comparatively high. Because complete misses were excluded from conditional distance summaries, these metrics must be interpreted together with Dice and the reported finite-case denominators. This reporting choice avoids assigning arbitrary distance penalties while preserving visibility of detection failures.

The external UMD-style experiment was intentionally stringent. Orientation and label descriptions were matched as closely as possible, but institutional source, acquisition parameters, scanner platform, pathology, and case mix remained different. All four models deteriorated substantially, and the absolute volume errors were large. The result should not be interpreted as isolating a specific source of domain shift; rather, it demonstrates that matching orientation and annotation ontology was insufficient to preserve internal performance. The small sample size limits precise model ranking, but the consistent degradation supports external validation before reuse in a new uterine MRI population.

From a biomedical image-processing perspective, these findings caution against treating architecture ranking on a single dataset as evidence of general robustness. Dataset composition, preprocessing, annotation rules, and optimization strategy can alter both absolute performance and the magnitude of differences among models. Reproducible comparisons should therefore report the complete processing pipeline, use fixed case-level partitions, evaluate complementary overlap and boundary metrics, and include an external or shifted test set whenever possible.

The results have practical implications for future uterine segmentation studies. nnU-Net remains an appropriate reference system for new tasks, while MedNeXt is a strong manually configured convolutional alternative. Public datasets should be expanded to include broader scanner, orientation, and pathology diversity, and label harmonization should be documented explicitly. Future work should also evaluate calibration or uncertainty, adaptation strategies, and larger multi-institutional external cohorts rather than relying only on incremental backbone changes.

This study has limitations. The external UMD-style cohort contained only 12 cases, pathology labels were unavailable, and scanner-specific effects could not be separated. The comparison with nnU-Net was intentionally system-level and does not isolate individual adaptive components. Finally, one expert radiologist generated the institutional annotations, so inter-reader variability was not measured.

In conclusion, segmentation robustness across uterine MRI depended on the clinical task, architectural family, pipeline configuration, and evaluation domain. MedNeXt was the strongest manually configured architecture, whereas nnU-Net remained the most reliable complete system. Marked external degradation across all models indicates that high internal accuracy is not sufficient evidence of transportability. For heterogeneous biomedical imaging applications, dataset-adaptive configuration, transparent evaluation, and external validation may be more consequential than selecting a nominally newer backbone alone.

## Data and code availability

The Uterine Myoma MRI Dataset (UMD) is publicly available through its original publication. The endometrial cancer and uterine mass datasets are not publicly available because of patient privacy and institutional data-governance restrictions but may be available from the corresponding author upon reasonable request, subject to appropriate ethical approvals and data-use agreements. Training, inference, evaluation scripts, environment specifications, and task-specific run examples are publicly available at https://github.com/AIPHL-McGill/UterusSegBenchmark.

## Acknowledgements

This manuscript was informed by the design, scope, and ongoing development of the uterine mass study. The authors gratefully acknowledge the radiologists, clinicians, residents, fellows, research coordinators and collaborating investigators whose collective efforts in data collection, quality assurance and stewardship have established and continue to maintain the uterine mass dataset. Their sustained contributions have created a valuable collaborative research resource that informed this comparative evaluation. The views, interpretations and conclusions presented are those of the authors and should not be interpreted as representing the views of the uterine mass collaborators or funding organizations. We also acknowledge the investigators responsible for the public release of the Uterine Myoma MRI Dataset, which enabled open evaluation in this domain.

## Funding

The uterine mass dataset was developed and is maintained through funding from the Fondation de l’Association des radiologistes du Québec (FARQ) and the Cancer Research Society (CRS). No specific funding was received for the preparation of this manuscript. Dr. James Man Git Tsui was supported by the Fonds de recherche du Québec – https://doi.org/10.69777/368850. Dr. Jérémy Dana is supported by the *Fonds de Recherche du Québec* (FRQ) Junior 1 Salary Award (#380243).

## Declaration of competing interest

The authors declare that they have no known competing financial interests or personal relationships that could have appeared to influence the work reported in this paper.

## Notes

### Competing Interest Statement

The authors have declared no competing interest.

### Author Declarations

This retrospective study were conducted under McGill University Health Centre Research Ethics Board approval. All institutional imaging and annotations used for this study were handled in accordance with this approval and applicable local data-governance requirements.

### Summary of Updates

This version revises the title and reframes the study around robustness and generalization of deep learning segmentation across heterogeneous uterine MRI tasks. The abstract, introduction, results interpretation, discussion, and conclusion were revised for clarity and consistency. Statistical reporting was updated to include Holm correction within each dataset. The nnU Net reference was updated to the peer reviewed publication. The study cohorts, experimental analyses, quantitative results, and main conclusions regarding model performance are unchanged.

